# The appendix orchestrates T-cell mediated immunosurveillance in colitis-associated cancer

**DOI:** 10.1101/2021.05.25.21257681

**Authors:** Maxime K. Collard, Julien Tourneur-Marsille, Mathieu Uzzan, Miguel Albuquerque, Maryline Roy, Anne Dumay, Jean-Noël Freund, Jean-Pierre Hugot, Nathalie Guedj, Xavier Treton, Yves Panis, Eric Ogier-Denis

**Author notes:** <u>Corresponding author</u>: Dr. Eric Ogier-Denis, INSERM 1149/1242, rue de la Bataille Flandres Dunkerque Bâtiment D 1^er^ étage 3 RENNES, France, **Email:**.

## Abstract

**Objective:** While appendectomy may reduce colorectal inflammation in patients with ulcerative colitis (UC), appendectomy has been suggested to be associated with an increased risk of colitis-associated cancer (CAC). The aim of this study was to explore the mechanism underlying the appendectomy-associated increased risk of CAC.

**Design:** Five-week-old male BALB/c mice underwent appendectomy, appendicitis induction or sham laparotomy. They were then exposed to azoxymethane/dextran sodium sulfate (AOM/DSS) to induce CAC. Mice were sacrificed 12 weeks later, and colons were taken for pathological analysis and immunohistochemistry (CD3 and CD8 staining). Human colonic tumors from 21 UC patients who underwent surgical resection for CAC were immunophenotyped and stratified according to the appendectomy status.

**Results:** While appendectomy significantly reduced colitis severity and increased CAC number, appendicitis induction without appendectomy led to opposite results. Intra-tumor CD3+ and CD8+ T-cell densities were lower after appendectomy and higher after appendicitis induction compared to the sham laparotomy group. Blocking lymphocyte trafficking to the colon with the anti-α4β7 integrin antibody or a sphingosine-1-phosphate receptor agonist suppressed the inducing effect of the appendectomy on tumors’ number and on CD3+/CD8+ intra-tumoral density. CD8+ or CD3+ T cells isolated from inflammatory neo-appendix and intravenously injected into AOM/DSS-treated recipient mice increased CD3+/CD8+ T-cell tumor infiltration and decreased tumor number. In UC patients with a history of appendectomy, intra-tumor CD3+ and CD8+ T-cell densities were decreased compared to UC patients without history of appendectomy.

**Conclusions:** In UC, appendectomy could suppress a major site of T-cell priming resulting in a less efficient CAC immunosurveillance.

**Significance of this study:** *What is already known on this subject?:*

- The protective effect of preemptive appendectomy is currently investigated as a therapy for refractory ulcerative colitis (UC), with encouraging results.
- An increased risk of developing colitis-associated cancer (CAC) caused by this promising treatment has been identified.
- Since it is commonly accepted that CAC is related to colitis severity and extent, this finding is counterintuitive and the mechanisms of this paradoxical effect remain unknown.

*What are the new findings?:*

- In a mouse model of CAC, less extended colitis associated with an increased number of tumors was observed. Intra-tumor T-cell infiltration was significantly reduced after appendectomy. Blocking lymphocyte trafficking to the colon with current or experimental UC treatments mimicked the appendectomy-associated phenotype whereas neo-appendicitis or appendix-primed T-cell injection in recipient mice increased intra-tumor T-cell infiltration and strengthened protection against CAC.
- In UC patients with CAC, appendectomy was associated with a decreased intra-tumor T-cell infiltration.
- These findings suggest that, in UC, appendectomy could suppress a major site of T-cell priming in the colon, resulting in a reduced CAC immunosurveillance.

*How might it impact on clinical practice in the foreseeable future?:*

- This work emphasizes the fact that precautions will be necessary if appendectomy becomes an accepted therapeutic option for the treatment of refractory UC.
- Innovative cell-based therapies and immunotherapies, such as the administration of stimulated autologous appendicular T cells in patients with CAC are promising options.

## Introduction

In 1871, Charles Darwin has described the appendix as a rudimentary and useless vestige, that appeared as a consequence of a progressive shrinking of the cecum due to diet changes in our distant ancestors.[1] This historical conception has been refuted by a modern analysis of the evolutive history showing that the appendix has been positively selected among mammals for at least 80 million years and has made multiple independent appearances without any association with diet changes or cecum shrinking[2, 3], pointing out a potential benefit of this structure, the function of which is still almost unknown.[4]

An important question has been raised regarding whether removing the normal appendix is safe or not over time. This question is especially important given the recent improvements in the treatment of ulcerative colitis (UC). The incidence of this inflammatory bowel disease (IBD) can reach up to 465 cases per 100,000 inhabitants in developed countries [5] and it is characterized by chronic inflammation of the rectum and colon. Additionally, the risk of developing colorectal cancer (CRC), referred to as colitis-associated cancer (CAC), is increased in UC patients.[6] A history of appendicitis is rare in UC patients and a reduced incidence of UC has been observed in families with a history of appendicitis.[7] This suggests either a protective effect of appendectomy for appendicitis [8] or that appendicitis and UC involve alternative inflammatory responses.[9]

Interestingly, a history of appendectomy in subjects younger than 20 reduces the risk of developing UC in the general population, only in case of actual appendix inflammation.[8, 10]. In line with these findings, the protective effect of preemptive appendectomy has been investigated as a potential therapy for refractory UC and a clinical improvement has been shown in up to 50% of patients.[11] The possible effect of appendectomy on UC clinical outcomes could be a promising therapeutic option [12] but so far, evidence is lacking to use elective appendectomy as a routine therapeutic procedure in UC.

Data on the use of appendectomy as a therapeutic option are contradictory. While some studies have shown a decreased colectomy rate after appendectomy in UC patients, especially when performed after the onset of the disease,[13] other studies have suggested an increased risk of CAC after appendectomy and no significant change in colectomy rate despite a significant reduction in colorectal inflammation.[14, 15] Since it is commonly accepted that CAC is related to colitis severity and extent,[16] this last finding is counterintuitive and the mechanisms of this paradoxical effect remain to be investigated.

We have previously shown that appendectomy performed in a mouse model of UC led to a spontaneous onset of colonic tumors.[15] To further decipher the underlying mechanisms involved in appendectomy-induced CAC tumorigenesis, we investigated appendectomy consequences on immunosurveillance and lymphocyte trafficking in a mouse model of CAC. We then confirmed these results in colorectal tumors from UC patients.

## Materials and methods

### Animal models

All experimental procedures were approved by our local Animal Ethics Committee and the French Ministry of Research in accordance with the European legislation (APAFIS n° 14004-2018030914101923v5 and 24604-2020030518127896v3).

Five-week-old male BALB/c mice were purchased from Charles River and housed in ventilated cages with free access to water and food under controlled temperature (22 ± 2°C) and humidity (50 ± 10%). Mice were randomly assigned to experimental and control groups and the azoxymethane/dextran sodium sulfate (AOM/DSS) model of CAC was induced.[17] Mice were injected intraperitoneally with AOM (10 mg/kg of body weight; Sigma-Aldrich) at day 0. One week later, mice were treated with DSS (MW=36,000–50,000; MP Biomedicals) in sterile drinking water according to the following sequence: 1.5% DSS for 7 days (first DSS cycle), sterile drinking water for 7 days, 1.5% DSS for 7 days (second cycle), sterile drinking water for 14 days and 1.5% DSS for 4 days (third cycle). Between the end of the third DSS cycle and sacrifice, mice had free access to DSS-free drinking water. Mice were weighed once a week all along the AOM/DSS protocol and the clinical severity of colitis was assessed during each DSS cycle based on the percentage of weight change. Mice were sacrificed 12 weeks after AOM injection. To induce chronic colitis without CAC, mice underwent 3 DSS cycles without prior injection of AOM (DSS-only protocol). Colonic specimens were collected for blinded histological evaluation and further biological and biochemical analyzes.

### Surgical procedures

Surgery was performed under general anesthesia (intraperitoneal injection of buprenorphine at 0.1 mg/kg and inhalation of 3% isoflurane during the induction, and then 1.5% isoflurane during the procedure). Three different surgical procedures were performed: appendectomy, appendicitis induction and sham laparotomy (thereafter referred to as “control”). All mice underwent a single surgical procedure. After initiation of anesthesia, the skin was shaved and prepped with 70% ethanol. In the left iliac fossa, 1-cm paramedian laparotomy was performed, and the cecum was externalized from the peritoneal cavity. The cecal patch was identified as a 2-mm white ovoid structure on the antimesenteric side of the cecum. This structure is a major lymphoid structure in the colon of mice, located at the end of the cecum, and corresponds to the human appendix lymphoid cellular structure.[15, 18] The appendectomy procedure consisted in the resection of the cecal patch. To do so, the cecal patch was suctioned with a 1-ml plastic syringe, ligated at its base with Corolene^®^ 8/0 thread and resected. Then, the cecum was reintegrated into the abdomen and the abdominal wall was closed with two layers (muscles and skin) of Filapeau^®^ 6/0 continuous sutures. To induce experimental appendicitis, the cecal patch was ligated at its base but not resected, leading to local inflammation that resolved spontaneously without any antibiotics or secondary appendectomy. For the sham procedure, the cecum was externalized and reintegrated into the abdomen without any ligation of the cecal patch (control group). Buprenorphine (0.1 mg/kg) could be administered postoperatively in case of apparent pain but no other treatments such as anti-inflammatory drugs or antibiotics were administered.

As part of the AOM/DSS protocol, surgery was performed at the same time as the AOM injection, i.e., 1 week before starting the first DSS cycle. AOM was injected into the peritoneal cavity at the end of the procedure, after abdomen closure. Similarly, as part of the DSS-only protocol, surgery was performed 1 week before starting the first DSS cycle. A third protocol called “surgery-only” was used to assess the effect of surgery in the absence of colitis and CAC. As part of this protocol, mice underwent surgery (appendectomy, appendicitis or sham laparotomy) and were sacrificed 1 week later without any administration of AOM or DSS.

### Human samples

From January 2006 to December 2017, all UC patients (n=21) who underwent surgical resection for CAC in our institution (Beaujon hospital, Clichy, France) were included. Patients with Crohn’s disease or unclassified IBD or with dysplastic lesions only were excluded. Patients’ clinical characteristics were retrospectively collected. Paraffin-embedded CAC samples were collected for pathological analysis and immunohistochemistry. Any history of appendectomy was obtained from the pathology report of (sub-)total colectomy or coloproctectomy describing the presence or the absence of an appendix on the surgical specimen. Paraffin-embedded blocks with colorectal tumor fragments were collected for immunohistochemistry.

This study was approved by our local Ethics Committee and the French Ministry of Research (n° 12.739) in accordance with the European legislation.

Detailed methods about experimental procedure and statistics are outlined in the supplementary materials.

## Results

### Appendectomy increases tumorigenesis of CAC without worsening colitis

We first investigated the impact of appendectomy on CAC development in the AOM/DSS model (figure 1A). A significantly increased number of colorectal tumors was macroscopically observed in the appendectomy group (24.5 [20.0-31.8] tumors) compared to the control group (sham laparotomy) (15.0 [11.8-22.3] tumors, P=0.0028, figure 1B-C). This increase was confirmed microscopically (13.5 [12.3-19.8] tumors *versus* 9.0 [7.5-10.3] tumors per slide, P <0.0001, figure 1D-E).

**Figure 1:**
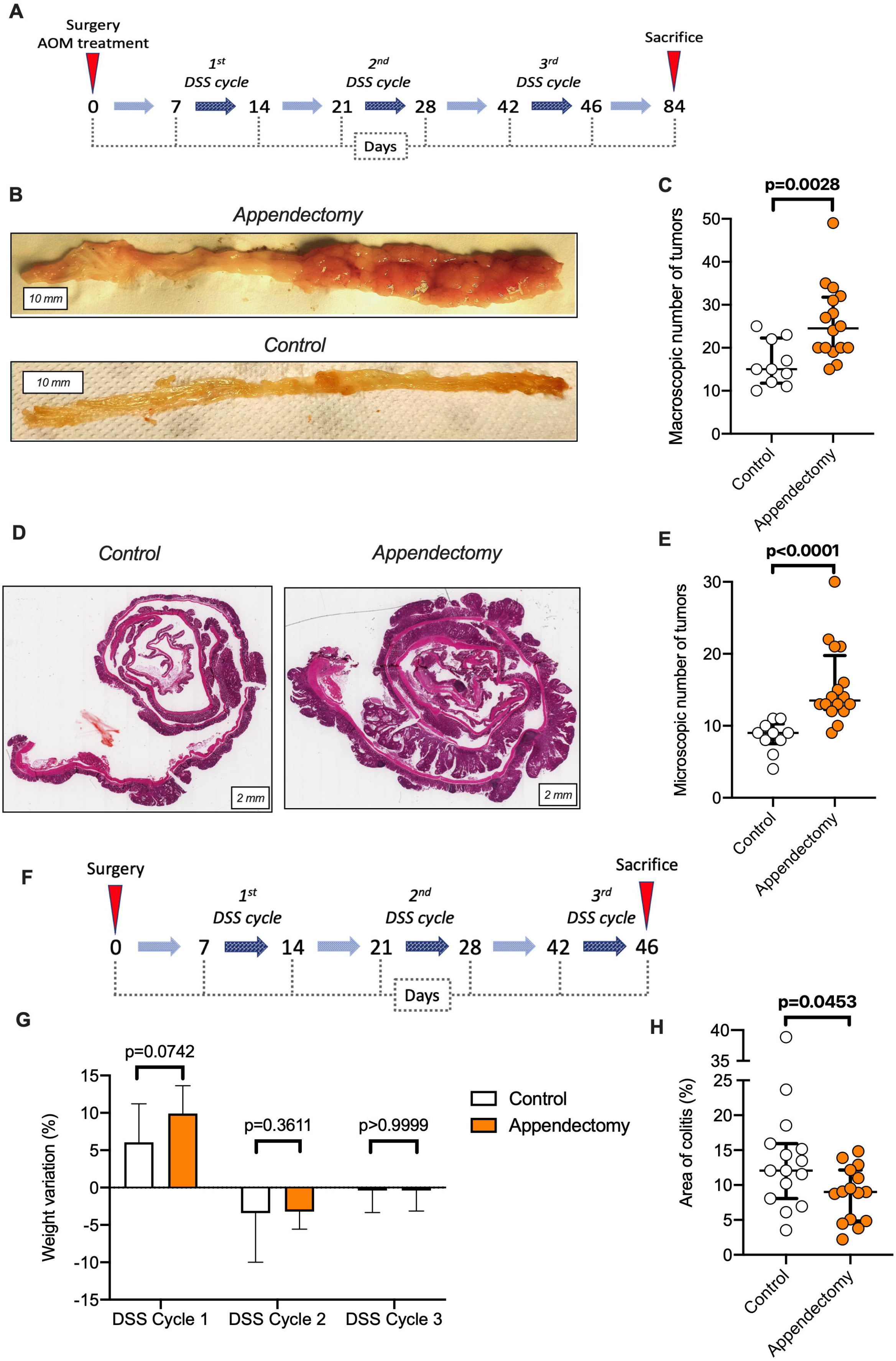
Appendectomy increases tumorigenesis of CAC and reduces colitis severity. Experimental schema of the AOM/DSS protocol used to induce CAC in mice. At the end of the AOM/DSS protocol (12 weeks after surgery and AOM injection), the entire colon from each mouse was removed and open longitudinally. (B) Representative picture of an opened colon for each group (distal part of the colon on the right side). (C) Tumor quantification in the appendectomy (n=16) and control (n=10) groups (macroscopic examination). Colons were fixed and embedded in paraffin and prepared as “Swiss rolls”. (D) Representative histological picture of a paraffin-embedded section stained with H&E reagent. (E) The number of tumors (microscopic examination) was counted in both groups. (F) Experimental schema of the DSS-only protocol. (G) Percentage of body weight change during each DSS cycle of the DSS-only protocol observed in the appendectomy (n=15) and control (n=15) groups. At the end of the DSS-only protocol, the entire colon was taken and stained with H&E reagent. Aperio ImageScope software was used to calculate the percentage of inflamed colonic epithelium surface within the entire colonic epithelium for each mouse. (H) Comparison of colitis extent after the DSS-only protocol between the appendectomy and control groups. In all boxplots, the error bars represent the 25^th^, 50^th^ (median) and 75^th^ interquartile ranges. Comparisons of 2 groups were performed using the Mann-Whitney test with 2-tailed p-value. A p-value <0.05 was considered statistically significant.

As chronic inflammation is the primary cause of CAC, we explored the impact of appendectomy on colitis severity after the third DSS cycle (DSS-only protocol, figure 1F). Body weight changes during each DSS cycle did not significantly differ between the appendectomy and control groups (figure 1G). However, the histological assessment revealed that appendectomy significantly reduced the extent of colitis compared to controls (P=0.0453, figure 1H), suggesting that appendectomy could moderately decrease the severity of chronic inflammation.

Reduced colonic inflammation following appendectomy has also been reported in UC patients.[11] However, it is classically admitted that the risk of developing CAC correlates with colitis severity in UC while we observed a concomitant paradoxical increase in the number of colonic tumors. To explore this counterintuitive result, mice were exposed to the AOM/DSS protocol at four different DSS concentrations from 0.5% to 2.0%. We found a positive correlation between the DSS concentration and colitis severity, and the number of colonic tumors (supplementary figure S1), confirming a role of inflammation in tumorigenesis and thus suggesting that appendectomy decreased tumorigenesis through a mechanism other than inflammation.

To assess whether appendectomy induced specific tumorigenic pathways, we profiled the molecular landscape of tumors from appendectomized and control mice. The comparative transcriptome analysis of tumors revealed a similar gene expression profile between both groups. Among the 23,517 RNA transcripts analyzed, only 27 (0.1%) were differentially expressed due to appendectomy (figure 2A). In addition, tumor proliferation assessed by immunohistochemistry staining of PCNA was similar in both groups (figure 2B). Thus, no specific tumor molecular pattern was identified in appendectomized mice.

**Figure 2:**
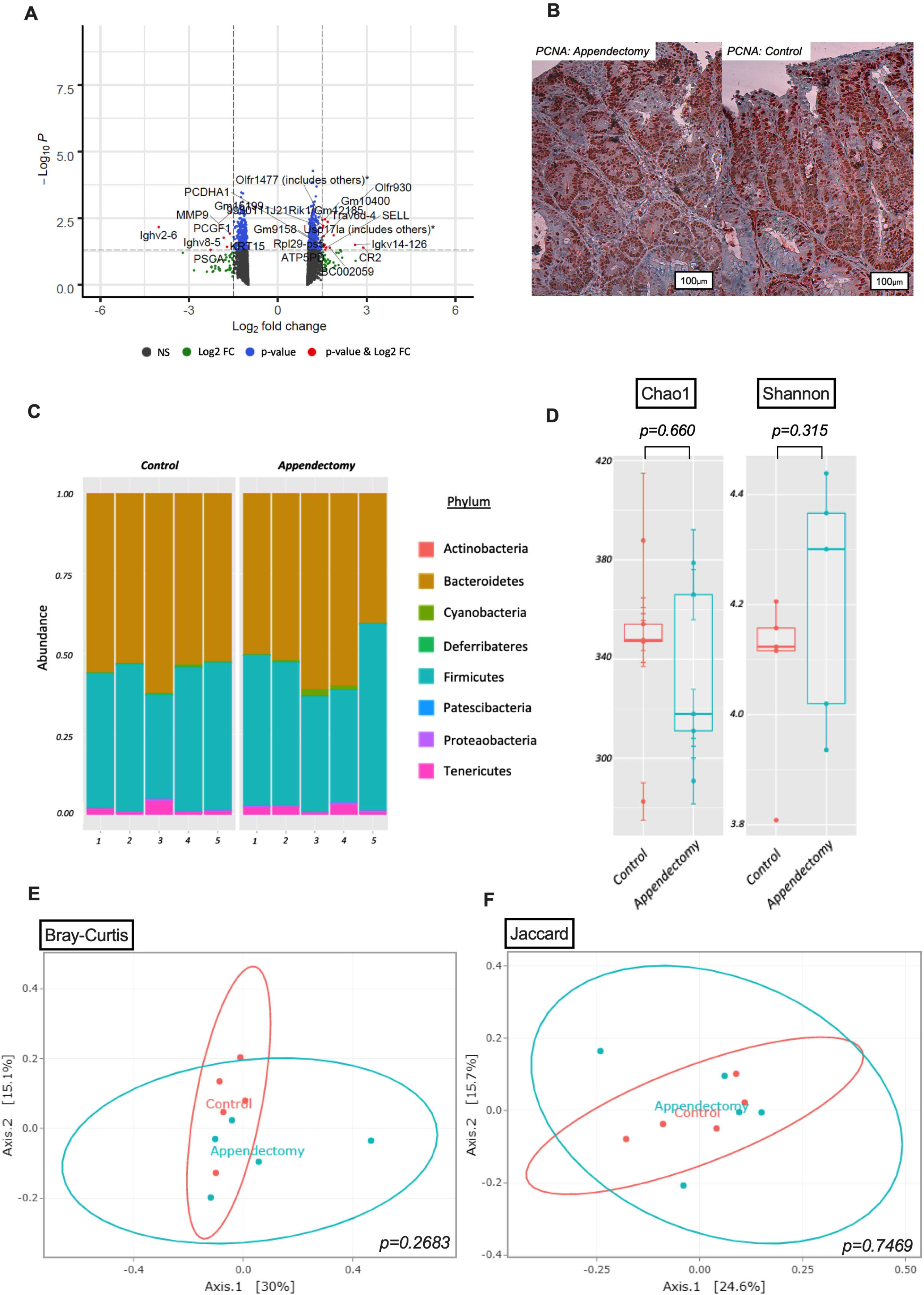
Transcriptome analysis of tumors, and assessment of tumor proliferation and fecal microbiota after appendectomy. To perform a transcriptome analysis of CAC, 11 mice were subjected to the AOM/DSS protocol with appendectomy (n=6) or sham laparotomy (n=5). Colonic tumors from each mouse were pooled and mRNA expression was analyzed by microarray using GeneChip® MouseGene2.0ST (Affymetrix). Only transcripts with a p-value <0.05 and an expression threshold >1.5 were considered differentially expressed between appendectomized and control mice. (A) Vulcano plot used in the differential gene expression analysis. The colored dots (green and red) represent the genes differentially expressed based on a p-value <0.05 (False Discovery Rate=2%; represented by a black hashed horizontal line) and a 1.5-fold expression difference (represented by two black hashed vertical lines). (B) Representative immunohistochemistry with the anti-PCNA antibody (Sc-56, Biotechnology, 1/100 dilution) of colonic tumors from control and appendectomized mice treated with AOM/DSS. (C) Relative abondance of the 8 most represented bacterial phyla in fecal samples of mice that underwent appendectomy (n=5) or sham laparotomy (n=5). The V4 variable region of the 16S rRNA genes was amplified by PCR in each sample and sequenced. The taxonomy of each filtered sequence was assigned using the 16S rRNA database Silva 132 Pintail 100. (D) Comparison of fecal microbiota alpha diversity assessed for richness (Chao1) and diversity (Shannon) between the appendectomy and control groups. Comparison of beta diversity according to the abundance determined using (E) the Bray-Curtis or (F) the Jaccard index between both groups. A p-value <0.05 was considered statistically significant.

The appendix may be instrumental in maintaining the intestinal microbiome [19] and gut dysbiosis could be a relevant hypothesis to explain the increased tumorigenesis.

The non-spore-forming Gram-negative bacteria, *Fusobacterium nucleatum*, have been associated with both acute appendicitis,[20] mild colitis in UC,[21] and more severe forms of CRC.[22] We assessed the intra-tumor infiltration of *Fusobacterium nucleatum* by qPCR in individual tumors. *Fusobacterium nucleatum* was detected in 1 out of 7 and in 1 out of 5 mice (supplementary figures S2A and S2B) in the appendectomy and control groups, respectively, indicating that *Fusobacterium nucleatum* was not directly involved in the tumorigenic mechanisms.

We further investigated the overall composition of the fecal microbiota one week after surgery and before DSS treatment. This timepoint was selected to identify the specific effects of appendectomy while avoiding the indirect effects of colitis and tumorigenesis on microbiota composition. 16s rRNA sequencing of the fecal microbiota revealed a similar abundance of the 8 most represented bacterial phyla between control and appendectomized mice (figure 2C). Both alpha diversity, assessed as the observed richness (Chao1) and diversity (Shannon), (figure 2D) and beta diversity, measured based on the Bray-Curtis or Jaccard indexes, did not significantly differ between both groups (figures 2E-F). Thus, the gut microbiota did not appear to play a major role in tumorigenesis induced by appendectomy.

### Appendectomy reduces intra-tumor T-cell infiltration

To determine the importance of the immunological function of the cecal patch (the murine equivalent of the lymphoid structures of the human appendix) in colitis-associated tumorigenesis, we focused on the anti-tumor immunity driven by CD3+ and CD8+ T cells.[23] Immunohistochemistry showed a significant decrease in CD3+ and CD8+ T-cell tumor infiltration after appendectomy (P=0.0001, figures 3A-B and P=0.031, figures 3C-D, respectively).

**Figure 3:**
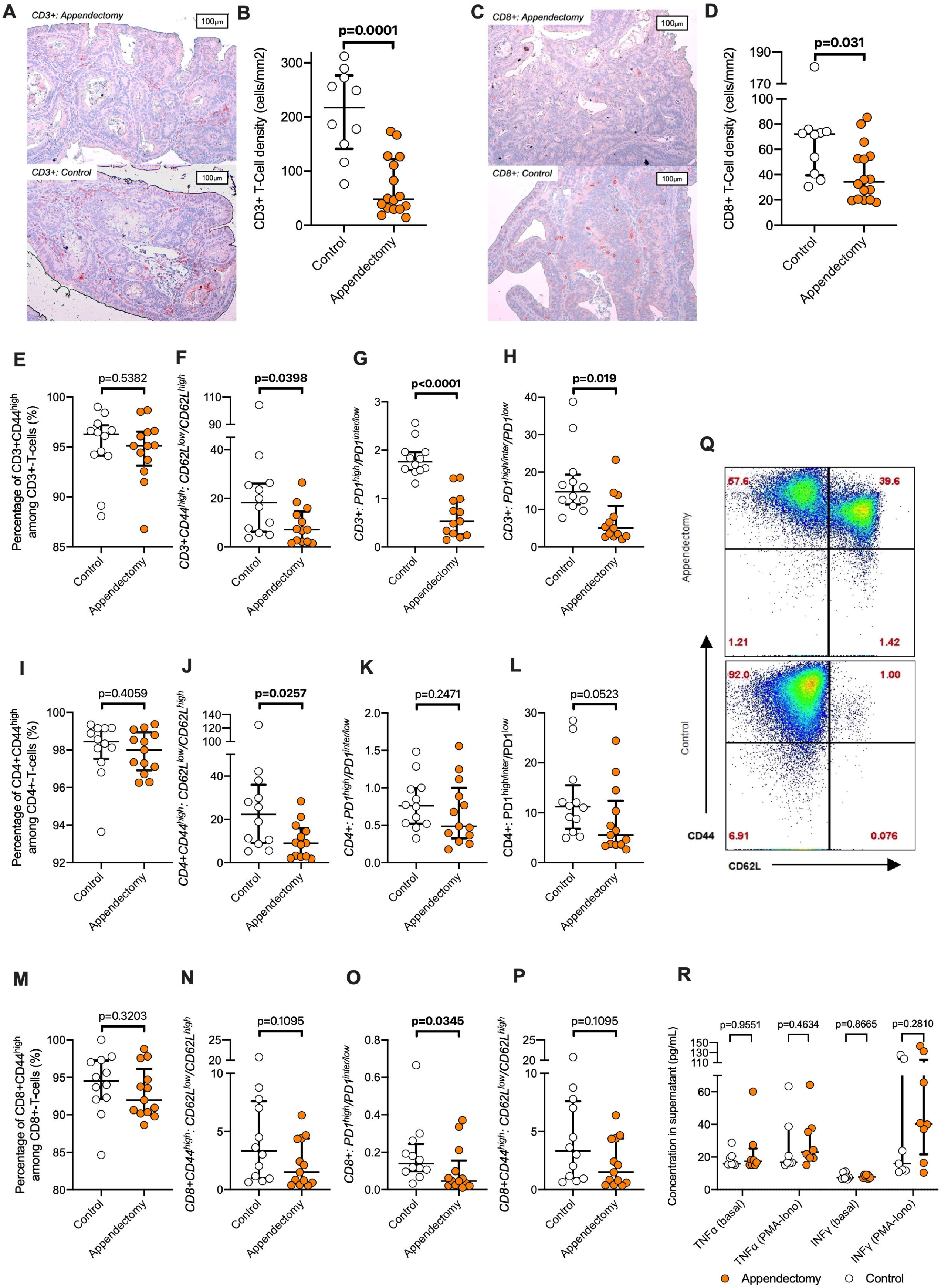
Appendectomy significantly alters intra-tumor T-cell immunity in mice. Paraffin-embedded sections of the colon from each mouse taken at the AOM/DSS protocol endpoint in the appendectomy (n=16) and control (n=10) groups were stained with anti-CD3 or anti-CD8 antibodies. (A) Representative CD3+ cell immunostaining of colonic tumors after appendectomy or sham laparotomy (control). (B) Quantification of intra-tumor CD3+ T-cell density by an automated observer-independent process using Aperio ImageScope software in the appendectomy and control groups. (C) Representative CD8+ immunostaining of colonic tumors after appendectomy or sham laparotomy (control). (D) Quantification of intra-tumor CD8+ T-cell density in the appendectomy and control groups. All colonic tumors from individual mice of the appendectomy (n=13) and control (n=12) groups subjected to the AOM/DSS protocol were resected and pooled. Intra-tumor CD3+ T-cells were isolated. Flow cytometry was performed to label CD3, CD4, CD8, CD62L, CD44 and PD1. Boxplots comparing intra-tumor cell labelling between the two groups in terms of (E) percentage of CD3+CD44^high^ cells among CD3+ cells, (F) CD62L^low^/CD62L^high^ ratio among CD3+CD44^high^ cells, (G) PD1^high^/PD1^inter/low^ ratio among CD3+ cells, (H) PD1^high/inter^/PD1^low^ ratio among CD3+ cells, (I) percentage of CD4+CD44^high^ cells among CD4+ cells, (J) CD62L^low^/CD62L^high^ ratio among CD4+CD44^high^ cells, (K) PD1^high^/PD1^inter/low^ ratio among CD4+ cells, (L) PD1^high/inter^/PD1^low^ ratio among CD4+ cells, (M) percentage of CD8+CD44^high^ cells among CD8+ cells, (N) CD62L^low^/CD62L^high^ ratio among CD8+CD44^high^ cells, (O) PD1^high^/PD1^inter/low^ ratio among CD8+ cells, (P) PD1^high/inter^/PD1^low^ ratio among CD8+ cells. (Q) Scatter plot of CD3+ cells isolated from appendectomy and control mice stained for CD62L (x-axis) and for CD44 (y-axis). The red number represents the percentage of labeled cells. (R) T cells were isolated from pooled colonic tumors of individual mice subjected to the AOM/DSS protocol after appendectomy (n=13) and sham laparotomy (n=12). 100,000 cells per mouse were stimulated or not with a cocktail of PMA-ionomycin. The boxplot represents the production of TNF-alpha and IFN-gamma by stimulated T cells from the appendectomy and control groups measured by ELISA. In all boxplots, the error bars represent the 25^th^, 50^th^ (median) and 75^th^ interquartile ranges. Comparisons of 2 groups were performed using the Mann-Whitney test with 2-tailed p-value. A p-value <0.05 was considered statistically significant.

We further characterized T cells infiltrating CAC by multiparameter flow cytometry (figures 3E-Q). The proportion of intra-tumor memory T cells (CD3+ and CD44^high^ cells among CD3+ cells) (figure 3E) was not affected by appendectomy (P=0.5382) even in the subgroups of CD4+ T cells (P=0.4059) (figure 3I) and CD8+ T cells (P=0.3203) (figure 3M). Among memory T cells (CD3+ and CD44^high^ cells), the ratio between effector (CD62L^low^) and central (CD62L^high^) memory T cells was significantly decreased after appendectomy (P <0.0001) (figures 3F and 3Q). Accordingly, it is noteworthy that the *SELL* mRNA encoding CD62L was overexpressed in tumors after appendectomy (figure 2A). Intra-tumor PD1^high^/PD1^inter/low^ and PD1^high/inter^/PD1^low^ T-cell ratios were also reduced in the appendectomy group, suggesting a lower proportion of intra-tumor exhausted T cells (figure 3G-H). However, the functional evaluation of intra-tumor T cells stimulated with PMA-ionomycin did not confirm this hypothesis (figure 3R).

Of note, at the DSS treatment endpoint (DSS-only protocol), CD3+ and CD8+ T-cell densities in the lamina propria were significantly reduced after appendectomy (supplementary figures S3 A-D), in line with the observation of less severe colitis. In contrast, one week after surgery without AOM/DSS treatment (Surgery-only protocol), CD3+ and CD8+ T-cell densities in the lamina propria were low and did not differ between groups (supplementary figure S3 E-H). The number of isolated lymphoid follicles in the colon was also unchanged (supplementary figure S3 I-J). These findings suggested that the immunological impact of appendectomy was mainly observed in case of colitis.

Overall, in tumors from appendectomized mice, CD8^+^ T-cell infiltration was decreased and the cell phenotype was switched from effector memory T cells to central memory T cells. This finding suggested that the increased CAC development after appendectomy was associated with decreased immune induction and surveillance.

### Inhibiting lymphocyte trafficking mitigates the pro-tumor effect of appendectomy

As a key inducer site, the appendix could orchestrate anti-tumor immunity in the colon in the context of chronic inflammation and antigenic stimulation. To test this hypothesis, intestinal homing of lymphocytes from the bloodstream to the gut was blocked with an anti-α4β7 integrin antibody in the one hand and lymphocyte egress from mesenteric lymph nodes and the appendix was inhibited with FTY720 on the other hand. Both blockades were supposed to mimic the pro-tumor effect of appendectomy.

As expected, anti-α4β7 integrin treatment eliminated the macroscopic and microscopic differences in tumor number between the control and appendectomy groups (figures 4A-B). Furthermore, each treatment independently reduced intra-tumor CD3+ and CD8+ T-cell densities that became similar between both groups (figures 4C-D). FTY720 treatment led to identical findings (figure 4E-H). Together, these experiments supported a defect of protective T-cell trafficking to the gut in the mechanism of appendectomy-associated tumorigenesis.

**Figure 4:**
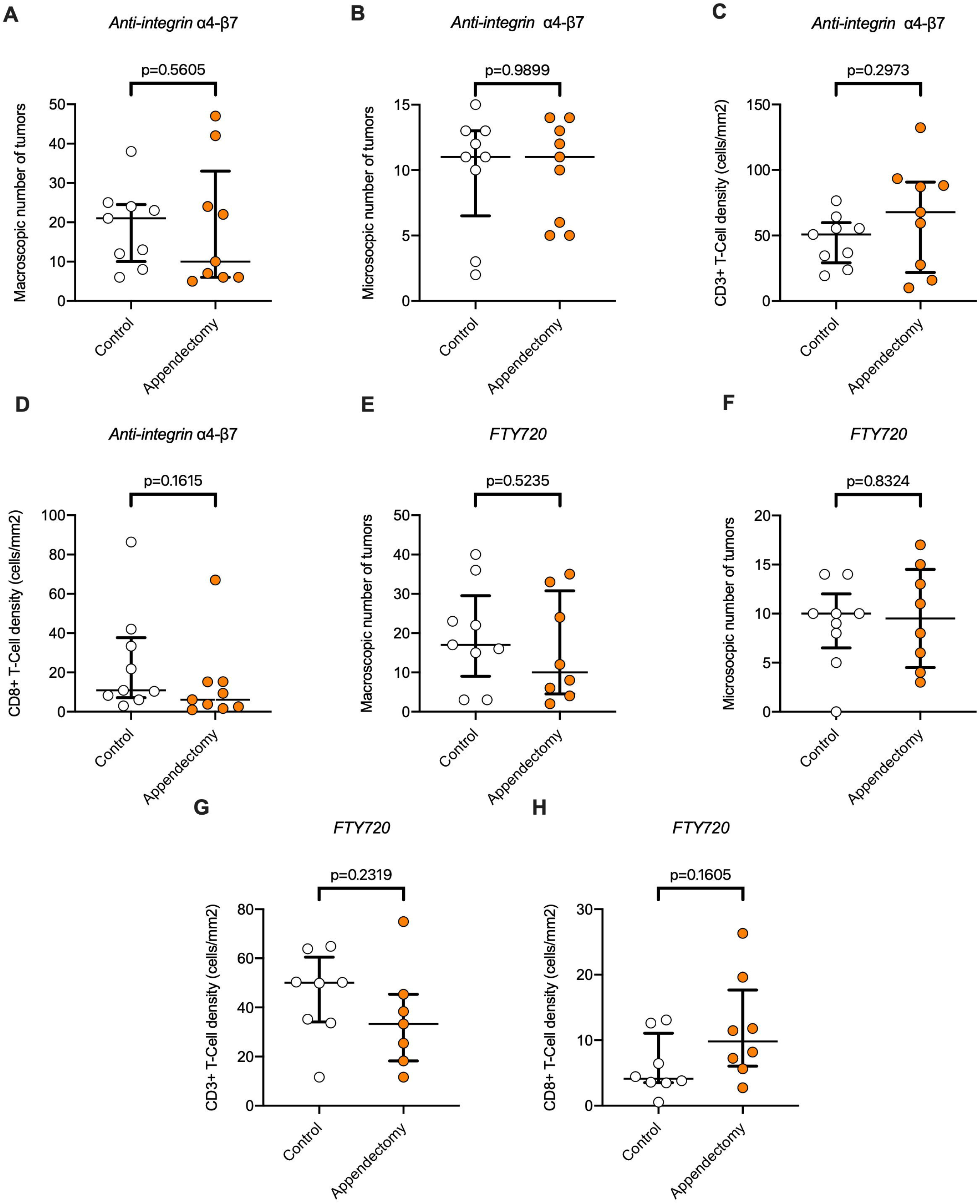
Blocking T-cell trafficking suppresses differences between the appendectomy and control groups. (A-D) To block T-cell trafficking to the colon, an anti-integrin α4-β7 antibody was administered twice a week (125 μg/100 μL of InVivoMAb anti-mouse LPAM-1, BE0034, Bioxcell per injection) from the first day of the first DSS cycle to the end of the AOM/DSS protocol in mice that underwent appendectomy (n=9) or sham laparotomy (n=9). At the AOM/DSS protocol endpoint, the colons were taken. Boxplots showing the comparisons between the appendectomy and sham groups in terms of (A) macroscopic number of tumors, (B) microscopic number of tumors, median intra-tumor CD3+ (C) and CD8+ (D) T-cell densities (immunohistochemistry). (E-H) To block T-cell trafficking to the colon, FTY720 (60 μg/100 μL per injection, SML0700, Sigma-Aldrich) was administered to 8 mice after appendectomy and to 9 mice after sham laparotomy following the AOM/DSS protocol and the frequency of injections was identical to that of the anti-integrin α4-β7 antibody. Boxplots showing the comparisons between the appendectomy and sham groups in terms of (E) macroscopic number of tumors, (F) microscopic number of tumors, median intra-tumor CD3+ (G) and CD8+ (H) T-cell densities (immunohistochemistry). In all boxplots, the error bars represent the 25^th^, 50^th^ (median) and 75^th^ interquartile ranges. Comparisons of 2 groups were performed using the Mann-Whitney test with 2-tailed p-value. A p-value <0.05 was considered statistically significant.

### Appendicitis protects against CAC and induces intra-tumor T-cell infiltration

We then assumed that appendicitis could have effects on tumorigenesis opposite to those induced by appendectomy, through the activation of immune surveillance and lymphocytes. Therefore, we explored the potential benefit of inducing neo-appendicitis on anti-tumor immune protection. As part of the AOM/DSS protocol, significant decreases in macroscopic (P=0.0075) and microscopic (P=0.0035) tumor numbers were detected in the appendicitis group compared to the control group (figures 5A-B). As expected, CD3+ and CD8+ T-cell infiltration was significantly increased in CAC after appendicitis (figures 5-C-D). Of note as part of the DSS-only protocol, no significant difference in body weight change after each DSS cycle was observed between the appendicitis and control groups (figure 5-E). However, the pathological analysis showed that appendicitis increased colitis extent throughout the colon compared to the control group (P=0.0453, figure 5-F). Thus, appendicular inflammation could worsen colitis and induce a phenotype opposite to that observed after appendectomy.

**Figure 5:**
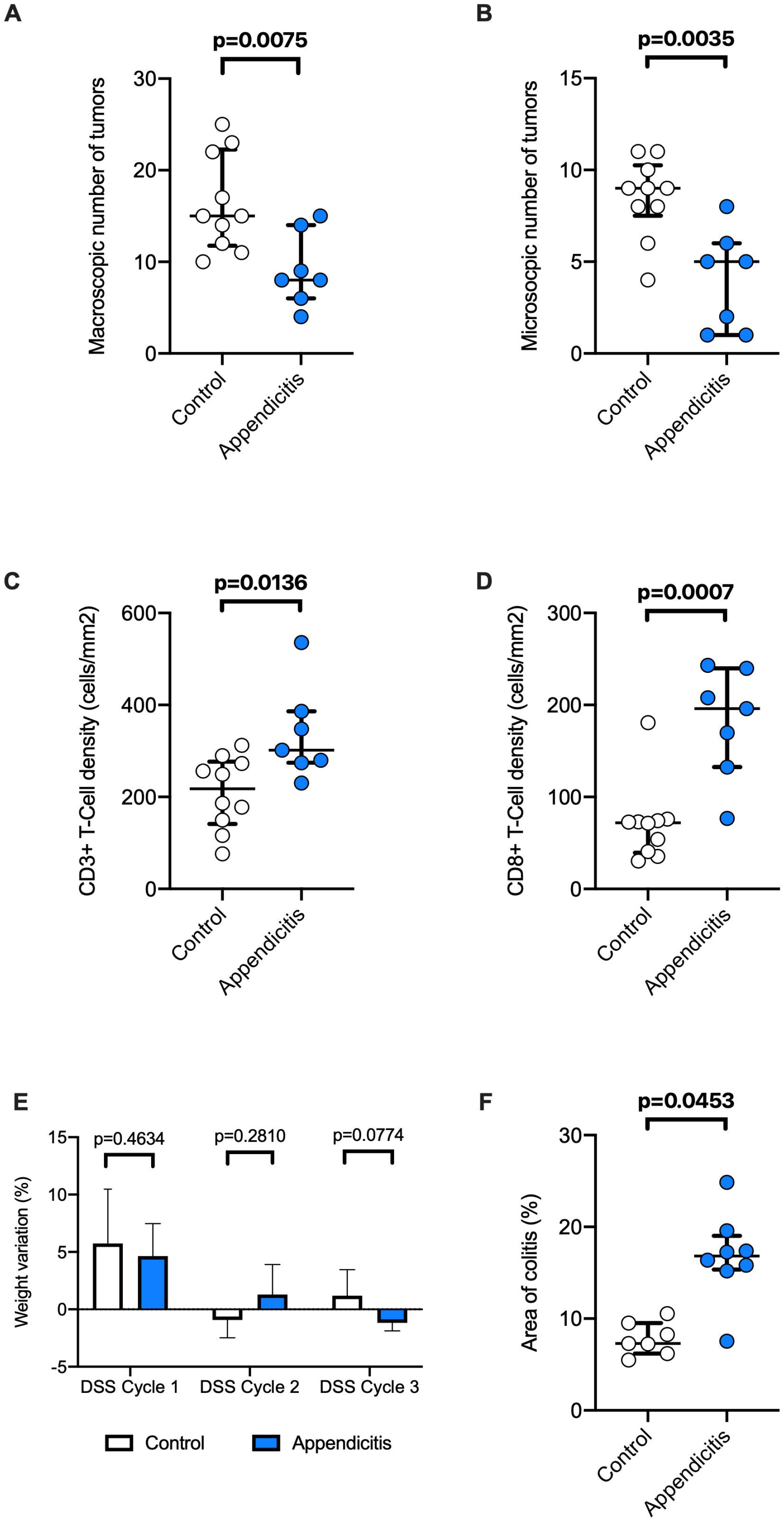
Appendicitis provides a partial protection against CAC and increases intra-tumor T-cell infiltration. Neo-appendicitis was surgically induced in 10 mice without appendectomy and sham laparotomy was performed in 7 mice. These two surgical procedures were performed at Day 0 of the AOM/DSS protocol and mice were sacrificed 12 weeks later. The colons were taken for the following analyzes. (A) Tumor quantification in the appendicitis and control groups. (B) Number of tumors visible on the H&E-stained slides (microscopic examination). Paraffin-embedded sections from each mouse were stained by immunohistochemistry (CD3 and CD8). Intra-tumor CD3+ (C) and CD8+ (D) T-cell densities. To assess the impact of appendicitis on colitis, appendicitis induction (n=8) or sham laparotomy (n=7) was performed in mice subjected to the DSS-only protocol. (E) Percentage of body weight change during each DSS cycle of the DSS-only protocol in the appendicitis and control groups. At the end of the DSS-only protocol, mice were sacrificed and their colon was taken and paraffin-embedded sections were stained with H&E reagent. The percentage of inflamed colonic epithelium surface within the entire colonic epithelium was calculated for each mouse. (F) Comparison of colitis extent after the DSS-only protocol between the appendicitis and control groups. In all boxplots, the error bars represent the 25^th^, 50^th^ (median) and 75^th^ interquartile ranges. Comparisons of 2 groups were performed using the Mann-Whitney test with 2-tailed p-value. A p-value <0.05 was considered statistically significant.

### Systemic injection of CD3+ or CD8+ T cells activated by appendicitis protects against CAC and induces intra-tumor T-cell infiltration

To further validate the role of circulating T cells, neo-appendicitis was surgically induced in donor mice and appendicular cells were isolated and purified one week later. T-cell depleted immune cells (CD45+ CD3-cells) or purified CD3+ T cells or CD8+ T cells were injected into the retro-orbital venous sinus of recipient mice that were further treated with AOM/DSS. Mice injected with CD3+ or CD8+ T cells showed a significant reduction in the number of colonic tumors compared to mice injected with CD45+ CD3-cells (figure 6A-B). A parallel increase in intra-tumor CD3+ and CD8+ T-cell densities was observed (figures 6C-D). No differences in tumor number or intra-tumor T-cell infiltration was noted between mice that received a CD3+ or CD8+ T-cell systemic injection.

**Figure 6:**
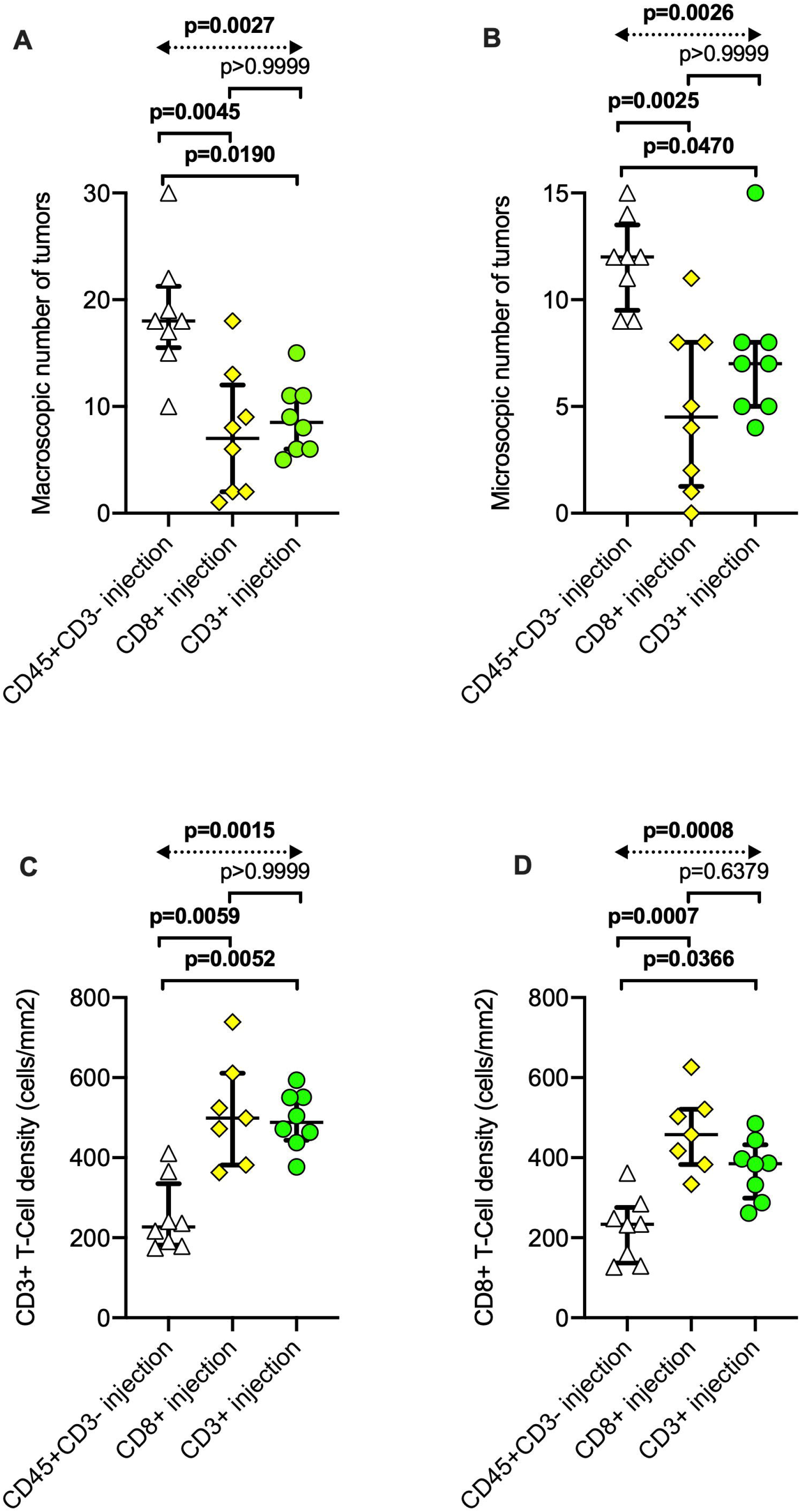
Injecting purified appendicular CD3+ or CD8+ T cells activated by appendicitis protects against CAC and increases intra-tumor T-cell infiltration. One week after the surgical induction of neo-appendicitis in mice, inflamed appendices were resected, and appendicular cells were isolated. CD8+ cells, CD3+ cells, and CD45+ cells depleted in CD3+ cells were purified. 5×10^5^ living filtrated CD8+ T cells were injected into 8 mice (CD8+ T-cell injection group), 5×10^5^ living filtrated CD3+ T cells were injected into 8 mice (CD3+ T-cell injection group) and 5×10^5^ living filtrated CD45+ cells depleted in CD3+ T cells were injected into 8 mice (CD45+ CD3-cell injection group). All mice were subjected to the AOM/DSS protocol. (A) Macroscopic quantification of colonic tumors in each group. (B) Microscopic quantification of colonic tumors from H&E-stained slides. Paraffin-embedded sections from each mouse were stained by immunohistochemistry (CD3 and CD8). Intra-tumor CD3+ (C) and CD8+ (D) T-cell densities quantified by an automated observer-independent process using Aperio ImageScope software. In all boxplots, the error bars represent the 25^th^, 50^th^ (median) and 75^th^ interquartile ranges. Comparisons of multiple groups were performed using the Kruskal-Wallis test and, only if the p-value was <0.05, multiple comparisons with post-hoc tests (Dunn’s test) were performed. A p-value <0.05 was considered statistically significant.

### The reduced intra-tumor CD3+ and CD8+ T-cell infiltration observed in mice after appendectomy is confirmed in CAC samples from UC patients

To confirm our results obtained in mice in humans, we analyzed colonic tumors from 21 consecutive UC patients who underwent colorectal resection for CAC between January 2006 and December 2017. Five patients (24%) had a history of appendectomy and 16 patients (76%) did not have any history of appendectomy. Clinical and oncological characteristics are summarized in table 1.

**Table 1.** Characteristics of patients with ulcerative colitis who underwent surgical resection for colitis-associated cancer.

|  | History of appendectomy | No history of appendectomy | p-value |
| --- | --- | --- | --- |
| **Population** | n= 5 | n= 16 |  |
| Gender (female vs. male) | 0 (0)<sup>1</sup>/5 (100) | 8 (50)/8 (50) | 0.111 |
| Age (years) | 38 (28-64)<sup>2</sup> | 49 (39-55) | 0.603 |
| Primary biliary cholangitis | 0 (0) | 4 (25) | 0.532 |
| Medical treatment for colitis in the 6 months before surgery | 3 (60) | 9 (56) | 1.000 |
| **Colorectal cancer** | n=6 | n=18 |  |
| Location: right or transverse colon *versus* left colon/rectum, | 3 (50)-2 (33)-1 (17)-0 (0) | 7 (39)-1 (6)-6 (33)-4 (22) | 0.197 |
| T-stage<sup>3</sup> |  |  | 0.539 |
| -*In situ* | 1 (17) | 1 (6) |  |
| -1 | 1 (17) | 3 (16) |  |
| -2 | 0 (0) | 2 (11) |  |
| -3 | 2 (33) | 10 (56) |  |
| -4 | 2 (33) | 2 (11) |  |
| N-stage: 0 / 1 or 2 | 4 (67)/2 (33) | 14 (78)/4 (22) | 0.618 |
| M-stage: 0 / 1 | 6 (100)/0 (0) | 18 (100)/0 (0) | 1.000 |

<sup>1</sup>number of patients (percent); <sup>2</sup>median (interquartile range); <sup>3</sup>according to the 8<sup>th</sup> TNM classification

More than one (synchronous tumors) CAC were found in the surgical specimen in one patient with a history of appendectomy (2 tumors) and in one patient without history of appendectomy (3 tumors). Thus, we assessed by immunohistochemistry 6 tumors in the appendectomy group and 18 tumors in the control group. The analysis of intra-tumor T cells confirmed that patients with a history of appendectomy had significantly lower CD3+ and CD8+ T-cell densities compared to patients without history of appendectomy (respectively P=0.0044, figures 7A-B and P=0.0472, figures 7C-D).

**Figure 7:**
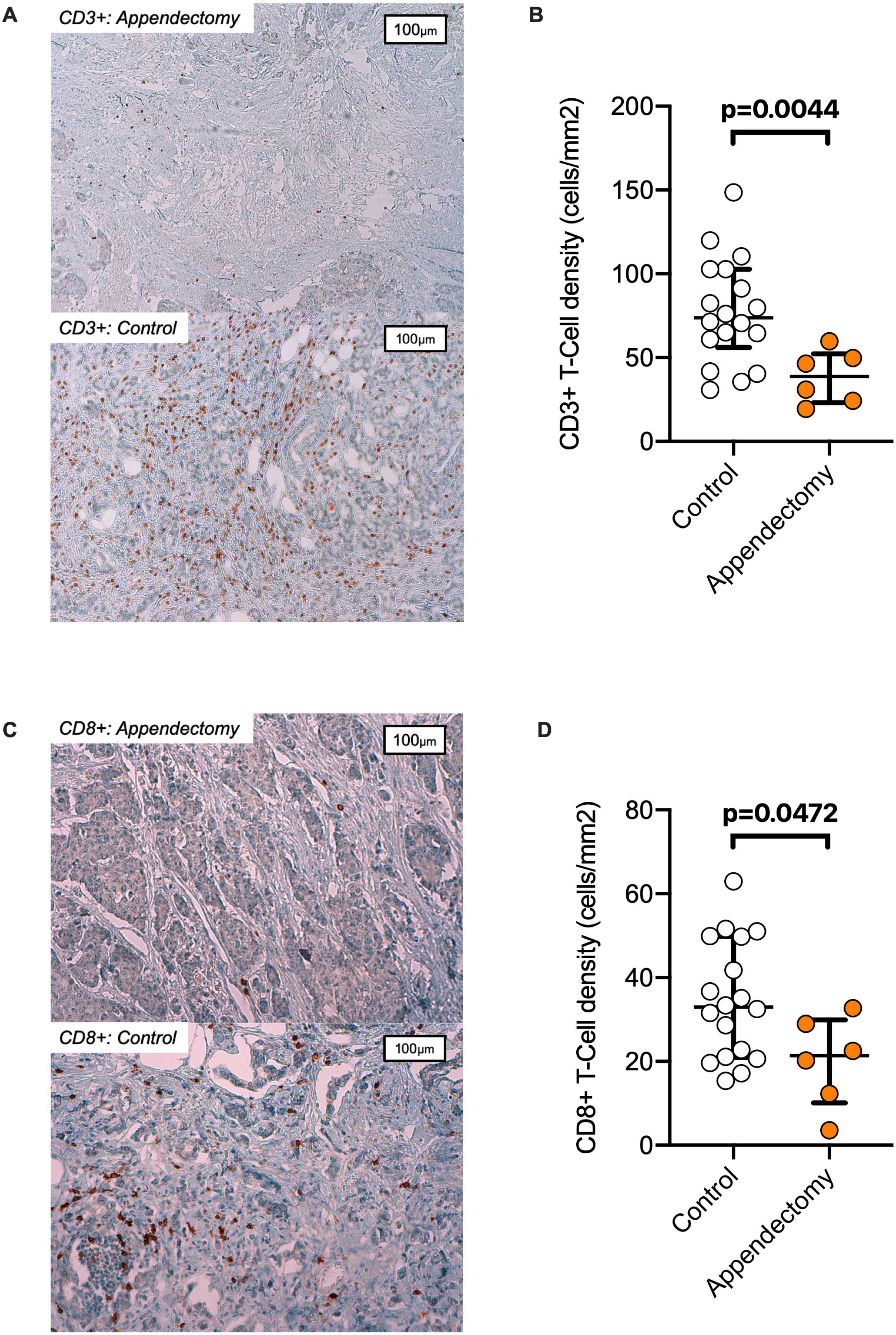
Appendectomy significantly alters intra-tumor T-cell immunity in human CAC. Tumors from patients with ulcerative colitis who underwent surgical resection for CAC were analyzed. All paraffin blocks embedding the colorectal tumors were collected to perform immunohistochemistry (CD3 and CD8). Of the 24 tumors analyzed, 6 tumors were from patients with a history of appendectomy and 18 tumors were from patients without history of appendectomy (control group). (A) Representative expression of CD3 in human colonic tumors from patients who underwent or not (control) appendectomy. (B) Quantification of intra-tumor CD3+ T-cell density in the appendectomy and control human tumors. (C) Representative expression of CD8 in human colonic tumors from patients who underwent or not (control) appendectomy. (D) Quantification of intra-tumor CD8+ T-cell density in the appendectomy and control groups. In all boxplots, the error bars represent the 25^th^, 50^th^ (median) and 75^th^ interquartile ranges. Comparisons of 2 groups were performed using the Mann-Whitney test with 2-tailed p-value. A p-value <0.05 was considered statistically significant.

## Discussion

Our results showed that appendectomy was associated with a reduced CD3+ and CD8+ T-cell infiltration in CAC in both mice and humans. In animal models, appendectomy increased colitis-associated tumorigenesis by preventing T-cell trafficking and the subsequent decrease in immune surveillance of cancer. As a mirror image, neo-appendicitis reinforced the protection against CAC through an increase in CD3+ and CD8+ T-cell immunosurveillance. The appendix thus appeared as a major inducer site for T-cell priming of colonic T lymphocytes and could also prime intra-tumor T cells in the context of CAC.

The negative impact of low intra-tumor CD3+ and CD8+ T-cell densities on the prognosis of sporadic CRC is well established.[24] The immunoscore based on CD3+ and CD8+ T-cell densities in sporadic colorectal tumors and in their invasive margins is a powerful prognostic marker for tumor evolution.[23] Intra-tumor CD3+ and CD8+ T-cell densities in CAC have been reported in two contradictory studies. Michael-Robinson *et al*. have highlighted higher densities in CAC,[25] whereas Seung Soh *et al*. have observed lower densities.[26] In both studies, low intra-tumor CD3+ and CD8+ densities were associated with a poorer tumor prognosis in CAC as in sporadic CRC.[26] Here, we found in a mouse model of CAC that lower densities of these cell populations were associated with a higher number of tumors, suggesting a protective role of intra-tumor CD3+ and CD8+ T cells.

After appendectomy, the number of CD3+ and CD8+ T cells was reduced in both humans and mice with CAC, raising the question of the role of the appendix in cancer immunosurveillance. Noteworthy, the lower intra-tumor CD3+ and CD8+ T-cell densities were associated with a change in T-cell phenotype characterized by a reduced CD62L^low^/CD62L^high^ ratio among memory T cells (CD3+ and CD44^high^). CD3+/CD44^high^/CD62L^low^ and CD3+/CD44^high^/CD62L^high^ cells are effector (Tem-cell) and central (Tcm-cell) memory T cells, respectively.[27] The proportion of Tem-cells and Tcm-cells generated after antigen presentation to naïve T cells is not constant and depends on the intensity of the antigen exposure: the differentiation into Tem-cells requires a strong antigen exposure while it is the opposite for Tcm-cells.[28, 29] The appendix is a lymphoid structure where intestinal microbiota and environmental antigens are sampled and presented to the immune system.[30] The appendix is thus a major priming site in the colon, able to locally induce Tem-cells and its surgical resection is likely to reduce T-cell education with a significantly decreased Tem-cell/Tcm-cell ratio.

Appendectomy is also associated with a decreased expression of PD1 by T cells in tumors. Intra-tumor exhausted T cells are characterized by a high PD1 expression and an impaired effector function.[31] Indeed, the capacity of these exhausted T cells to proliferate and produce effector cytokines is reduced. Here, we found a low proportion of PD1^high^ T cells after appendectomy but similar cytokine levels between the appendectomy and control groups. This finding suggests that PD1^high^ T cells are not exhausted T cells. Beswick et *al*. have shown that PD1 is upregulated in inflamed colonic mucosa.[32] In line with our results, Yassin et *al*. have shown a progressive increase in PD1 expression over time in the mucosal T-cell subset using the AOM/DSS model.[33] Further CAC treatment with antibodies directed against PD1 has failed to reactivate PD1^high^ T cells directed against the tumors.[33] Recently, we have shown that using anti-PD1 treatment in the AOM/DSS model led to an increased tumor proliferation.[34] Therefore, PD1 upregulation could reflect previous colonic inflammation rather than an exhausting state of intra-tumor T cells.

CAC is a consequence of DSS-induced colonic inflammation. In the DSS-only model of colitis, colitis extent was reduced and the CD3+ and CD8+ T-cell densities in the lamina propria were significantly reduced after appendectomy. These findings are consistent with a loss of T-cell priming in the appendix even if other mechanisms could be involved.[35, 36, 37]

T-cell trafficking blockade with an anti-α4β7 integrin antibody or FTY720 strongly reduced the median number of tumors in the AOM/DSS model of CAC. These drugs are effective against colonic inflammation,[38, 39] and they could thus indirectly reduce colitis-associated tumorigenesis. However, the loss of differences in tumor number and the reduced intra-tumor CD3+/CD8+ T-cell densities between the appendectomy and control groups after trafficking blockade suggested that the effect of appendectomy could be mediated by lymphocyte recirculation towards the colonic lamina propria and tumors. Finally, a protection against CAC induced by intra-tumor CD3+/CD8+ T cells primed in the appendix was strongly supported by the T-cell transfer experiments. Indeed, an intravenous injection of intra-appendicular CD3+/CD8+ T cells obtained from mice in which neo-appendicitis was induced increased tumor infiltration by T cells and limited tumor number.

If the appendix plays a crucial role in CAC, why is the oncological immunosurveillance provided by CD8+ T cells primed in the appendix not effective in sporadic CRC? In industrialized countries, about 10% of the population will undergo appendectomy during their life.[40] Despite a high incidence of sporadic CRC, a history of appendectomy is not known as a risk factor.[41] However, in most cases, appendectomy is performed for acute appendicitis. Indeed, a Taiwanese national cohort study has confirmed that a history of appendectomy for appendicitis was not associated with a risk of cancer (hazard ratio (HR): 1.02, 95%CI [0.90-1.16]) but a history of incidental appendectomy without appendicitis was significantly associated with a risk of sporadic CRC (HR: 2.90, 95%CI [2.24-3.75]) [42]. This finding suggests that the appendix could also play a role in sporadic CRC.

In conclusion, using a mouse model of CAC, we showed that appendectomy induced a pro-tumorigenic effect mediated by intra-tumor T-cell infiltration. Blocking cell egress from the appendix or T-cell homing to the colon mimicked the appendectomy-associated phenotype whereas re-injecting appendix-primed T cells increased intra-tumor T-cell infiltration. In UC patients with CAC, appendectomy was associated with a decreased intra-tumor T-cell infiltration. Our data open new perspectives for innovative cell-based therapies and immunotherapies, such as the administration of stimulated autologous appendicular T cells in CAC patients.

## Data Availability

Data are available on reasonable request.

## Acknowledgements

We thank Nathalie Colnot, Raphaelle Liquard, Ian Morilla, Olivier Thibaudeau and Laure Wingertsmann for their substantial contribution to this project.

## Data availability

Data are available on reasonable request.

