## Supplementary file for "The appendix orchestrates T-cell mediated immunosurveillance in colitis-associated cancer"

**Supplementary materials**

*Pathology*

After sacrifice of mice, the entire colon was removed, opened in the longitudinal axis and macroscopically-visible tumors were counted. Then, swiss rolls of colons were fixed for 24h in 10% formalin and embedded in paraffin to observe the full-length organ. Paraffin-embedded sections (5 µm) were deparaffinized, stained with H&E reagent and the number of microscopically-visible tumors was determined. To assess the histological extent of colitis, tissue sections from mice that underwent the DSS-only protocol were digitized (Scanscope AT turbo, Leica). Histological lesions of colitis were delineated with Aperio ImageScope software to calculate the percentage of inflamed colonic epithelium surface within the entire colonic epithelium for each mouse.

*Immunohistochemistry*

Paraffin-embedded colonic sections (5 µm) from each mouse were prepared for immunohistochemistry using antibodies directed against CD3 (ab16669, Abcam, 1/150 dilution) and CD8 (ab209775, Abcam, 1/1,000 dilution). Tissue slides from AOM/DSS-treated mice were also stained with antibodies directed against PCNA (Sc-56, Biotechnology, 1/100 dilution) to assess tumor cell proliferation. Tumor sections from human samples were analyzed by immunohistochemistry using antibodies directed against CD3 (A0452, Dako, 1/50 dilution) and CD8 (M7103, Dako, 1/50 dilution). Immunostained slides were digitized with Scanscope AT turbo, Leica. Using Aperio ImageScope software, human and mouse tumors were delineated. Tumor surfaces and the number of cells stained with the specific antibodies were automatically quantified. The median intra-tumor CD3+ and CD8+ T-cell densities were then calculated. This method was thus observer-independent. The lamina propria was also delineated on slides from mice that underwent the Surgery-only and DSS-only protocols. The surface and number of cells stained with the specific antibodies were quantified to assess the median CD3+ and CD8+ T-cell densities.

*Isolation of intra-tumor T cells*

Colonic tumors obtained at the end of the AOM/DSS protocol were collected, taking care to not remove the adjacent healthy colon. All tumors from the same colon were pooled in 10 mL of RPMI 1640 medium with GlutaMAX (61870-010, Gibco) supplemented with 10 mg of type IV Collagenase (LS004188 Serlabo), 0.5% of fetal bovine serum (FBS) and 10 mg of DNase (DN25-100MG Sigma). Fresh tumors were transferred to gentleMACS tubes (130-096-334, Miltenyi). Digestion was performed with gentleMACS Dissociator for 36 minutes at 37°C and with subsequent centrifugation at 930 rpm (37C_m_LIDK_1 program). After collagenase digestion and mechanical disruption, a single-cell suspension was obtained after filtration with 100-μm and then 40-μm cell strainers and washed twice with RPMI. To increase T-cell concentration, intra-tumor T cells were selected using Mouse CD90.2 MicroBeads (130-121-278, Miltenyi) according to the manufacturer’s instructions. Magnetic separation was performed with the MultiMACs Separator Plus (130-098-637, Miltenyi).

*Flow cytometry*

Cell suspensions were Fc-blocked (FcR Blocking mouse, 130-092-575 Miltenyi) and dead cells were stained with LIVE/DEAD Fixable Aqua Dead Cell Stain Kit-AmCyan (L34957, Thermofisher). Then, cells were incubated for 20 minutes at 4°C in the dark with a cocktail of antibodies directed against CD3 (APC-Vio770, 130-119-793 Miltenyi), CD4 (FITC, 130-118-692, Miltenyi), CD8 (PE-Vio770, 130-119-123, Miltenyi), CD62L (APC, 130-112-837, Miltenyi), CD44 (PE, 130-118-694, Miltenyi) and PD1 (PE-CF594, 562523 BD). Samples were washed twice with PBS (phosphate buffered saline). Samples were acquired using LSRFortessa (BD) and analyzed with FlowJo v10.

*T-cell stimulation and cytokine measurement by ELISA*

For each mouse, 100,000 intra-tumor T cells were transferred to 96-well plates with 200 μL of cell culture medium in the presence or in the absence of a stimulation cocktail of PMA and ionomycin (00-4970-93; 1/500 dilution; Thermofisher). Cell culture medium included RPMI 1640 with GlutaMAX, 10% of FBS and 1% of antibiotic-antimycotic (15240096, Thermofisher). After 16 hours in a humidified 37°C incubator with 5% CO_2_, supernatants were collected and stored at -20°C for cytokine assay. Concentrations of TNF-α and INF-γ were measured by ELISA according to the manufacturer’s instructions (respectively 88-7324-22 and 88-7314-22; Thermofisher). Detection ranges were 8-1,000 pg/mL for TNF-α and 15-2,000 pg/mL for INF-γ.

*Transcriptome analyzes*

Total RNAs were extracted from fresh colonic tumors using the RNAble Kit (Eurobio) and quantified with a nanodrop-1000 spectrophotometer (Thermofisher). Microarray processing was performed by a genomic platform (genom’IC, Cochin Institute, Paris, France). After validation of the RNA quality with the Bioanalyzer 2100 (using the Agilent RNA6000 nano chip kit), 100 ng of total RNAs were reverse transcribed using the GeneChip® WT Plus Reagent Kit (Thermofisher). Briefly, the resulting double-strand cDNA was used for *in vitro* transcription with T7 RNA polymerase (all these steps are included in the WT cDNA synthesis and amplification kit from Thermofisher). After purification according to Thermofisher protocol, 5.5 μg of Sens Target DNA were fragmented, biotin labelled and controlled using the Bioanalyzer 2100. cDNAs were then hybridized to GeneChip® MouseGene2.0ST (Affymetrix) at 45°C for 17 hours, and the chips were washed on the FS450 fluidics station (Affymetrix) and scanned using the GCS3000 7G. Scanned images were then analyzed with Expression Console software (Affymetrix) to obtain raw data (CEL files) and metrics for Quality Controls. No apparent outlier value was detected. CEL files were normalized by Robust Multi-array Averaging (RMA) in the Bioconductor R with the Brain Array custom CDF vs 23. Statistical analyzes were performed with Partek® GS. A t-test was used to explore differences in expressed genes between the appendectomy and control groups. Only genes with p-values <0.05 and expression fold-changes >1.5 were considered differentially expressed between both groups.

*Fecal microbiota characterization*

Mouse fecal samples were collected one week after surgery and frozen at -80°C. DNA was extracted using the QIAamp Fast DNA Stool Mini Kit (51604, Qiagen). A mechanical lysis with FastPrep (MP Biomedicals) was added to the protocol before thermal lysis. DNA concentration was measured using the Qubit dsDNA High Sensitivity Assay Kit (Q32851, Thermofisher) and adjusted for each sample to 5 ng/µL. 16S rRNA genes were amplified by PCR with universal primers amplifying the V4 variable region (515F: GTGCCAGCMGCCGCGGTAA and 806R: GGACTACHVGGGTWTCTAAT).[1] Barcodes and Illumina sequencing adapters were attached using the Nextera XT Index Kit (Illumina). Amplicons were purified using Agencourt AMPure (Beckman Coulter), quantified by qPCR using the KAPA Library Quantification Kit (Roche), pooled in equimolar concentration and diluted to 5.5 pM for sequencing. Sequencing was performed on the Illumina Miseq (600 cycles, 2x300 bp, paired sequences). Sequences were analyzed with Galaxy-supported pipeline, called FROGS (Find, Rapidly, OTUs with Galaxy Solution).[2] We assembled the paired readings, removed sequences not related to the two V4 primers, cut the primers, removed sequences shorter than 200 bp and longer than 500 bp and finally removed all sequences containing an ambiguous base. Sequence clustering was performed with a similarity threshold of 97% using the Swarm algorithm and chimeric sequences were detected with the UCHIME method. A filtering tool was used to remove parasitic clusters, with reading abundances <0.005% of the total number of readings. The taxonomy of each filtered sequence was assigned with the 16S rRNA database Silva 132 Pintail 100. Alpha and beta diversity measurements were performed with the FROGSSTAT Phyloseq script on the normalized abundance data. Alpha diversity indices (Chao1 and Shannon) were calculated for each sample and compared between the appendectomy and control groups using an ANOVA analysis. A principal coordinates analysis based on the distance matrix of the beta diversity indices (Bray Curtis and Jaccard) was used to visualize differences in microbial composition between groups. Significance was assessed using a PERMANOVA (Permutational Multivariate Analysis of Variance Using Distance Matrices) test. A P-value <0.05 was considered significant.

*Quantitative PCR for the detection of Fusobacterium nucleatum DNA*

Genomic DNA was extracted from fresh colonic tumors using the DNeasy Blood and Tissue kit (Qiagen). DNA was quantified using a Qubit 3.0 Fluorometer (Thermo Fisher Scientific). Real-time qPCR was performed with 10 ng or 40 ng of DNA sample, 10 μM of primers and MesaBlue qPCR MasterMix (Eurogentec). *Fusobacterium nucleatum* DNA was detected using the following primers: forward 5’-CCAACCATTACTTTAACTCTACCATGTTCA-3’ and reverse 5’-GTTGACTTTACAGAAGGAGATTATGTAAAAATC-3’.[3] To detect the presence of bacterial DNA in samples, a 16S rDNA non-specific PCR was performed using the primers U968 5’-GAACGCGAAGAACCTTAC-3’ and L1401 5’-GCGTGTGTACAAGACCC-3’.[4] PCR were carried out using a LightCycler 480 instrument (Roche Diagnostics). Initial denaturation was performed at 95°C for 10 min, followed by 45 cycles consisting of 95°C for 15s and 60°C for 45s. A dissociation step was added, and dissociation curves were analyzed to confirm amplification fidelity. Positive PCR products were sent for sequencing to Eurofins Genomics, and sequences were analyzed through BLAST program (NCBI) to confirm *Fusobacterium nucleatum-*specific amplification.

*Blocking lymphocyte trafficking*

An anti-integrin α4-β7 antibody and FTY720 (a sphingosine-1-phosphate receptor agonist) were independently used to limit lymphocyte trafficking to the colon. From Day 1 of the first DSS cycle (i.e., 1 week after appendectomy or sham surgery) until sacrifice, 125 μg/100μL of InVivoMAb anti-mouse LPAM-1 (integrin α4β7) (BE0034, Bioxcell) or 60 μg/100μL of FTY720 (SML0700, Sigma-Aldrich) were intraperitoneally administered twice a week.

*Transfers of systemic immune cells isolated from inflamed appendices*

One week after appendicitis induction, mice were sacrificed and inflamed appendices were resected and pooled in gentleMACS tubes (130-096-334, Miltenyi) containing 10 mL of RPMI 1640 with GlutaMAX (61870-010, Gibco) supplemented with 10 mg of type IV Collagenase (LS004188 Serlabo), 0.5% of FBS and 10 mg of DNase (DN25-100MG Sigma). Cell digestion and dissociation were performed using the gentleMACS Dissociator for 36 minutes at 37°C with subsequent centrifugation at 930 rpm. After incubation, the single-cell suspension was obtained after filtration with 100-μm and 40-μm cell strainers and washed twice with RPMI. At this step, 2.1x10^8^ living cells were obtained and then divided into two different tubes (tubes A and B). Tube A was used to isolate CD8+ T cells from inflamed appendices using the Mouse CD8a+ T-cell Isolation Kit (130-104-075, Miltenyi). Magnetic separation was performed using the MultiMACs Separator Plus (130-098-637, Miltenyi) according to the manufacturer’s instructions. Tube B was used to isolate CD3+ T cells using Mouse CD90.2 MicroBeads (130-121-278, Miltenyi). Appendicular cells depleted in CD3+ T cells were then incubated with Mouse CD45 MicroBeads (130-052-301, Miltenyi). Cell isolation quality was controlled by flow cytometry. Among CD8+ filtered T cells, 87% of living cells were CD8+ and 0.007% were CD4+. Among CD3+ filtered T cells, 80% of living cells were CD3+, 38% were CD4+ and 39% were CD8+. Among CD45+ cells depleted in CD3+ T cells, 99% of living cells were CD45+ and 92% were CD3-.

CD8+ filtered T cells (CD8+ T-cell injection group), CD3+ filtered T cells (CD3+ T-cell injection group) and CD45+ cells depleted in CD3+ T cells (CD45+CD3- cell injection group) were respectively injected into 8 mice. Each mouse received a standardized injection containing 5x10^5^ living cells in 100 µL of PBS in the retro-orbital venous sinus. At the same time as these systemic cell injections, mice received an intraperitoneal injection of AOM and were then exposed to the 3 DSS cycles according to the AOM/DSS protocol. After sacrifice, 12 weeks after the beginning of the AOM/DSS protocol, tumors were quantified macroscopically and microscopically, and intra-tumor CD3+ and CD8+ T-cell densities were assessed after immunohistochemistry as described above.

*Statistics*

Quantitative values are expressed as a median [25^th^ percentile-75^th^ percentile]. Comparisons of 2 groups were performed using the Mann-Whitney test with 2-tailed P-value. Comparisons of multiple groups were performed using the Kruskal-Wallis test and, only if the P-value was <0.05, multiple comparisons with *post hoc* tests (Dunn’s test) were performed. A P-value <0.05 was considered statistically significant (2-sided tests). Statistical analyzes were performed using Prism v8.2.1 (GraphPad Software).

**Supplementary figures**

**Supplementary Figure S1:** Relationship between the severity of DSS-induced colitis and the number of colonic tumors.

Four groups of mice were subjected to the AOM/DSS protocol without surgical intervention and treated with different DSS concentrations: 0.5% (n=9), 1.0% (n=10), 1.5% (n=10) and 2.0% (n=9). Mice were sacrificed 12 weeks after AOM injection. (A) Percentage of body weight change during each DSS cycle in the 4 groups. (B) Macroscopic quantification of colonic tumors in each group. (C) Microscopic quantification of colonic tumors from H&E-stained slides. In all boxplots, the error bars represent the 25^th^, 50^th^ (median) and 75^th^ interquartile ranges. Comparisons of multiple groups were performed using the Kruskal-Wallis test. A p-value <0.05 was considered statistically significant.

**Supplementary figure S2:** *Fusobacterium nucleatum* intra-tumor infiltration is not influenced by appendectomy.

(A) Result of double PCR using *Fusobacterium*-specific primers in tumor DNA samples. M, DNA ladder; lanes 1-16, PCR products from tumor DNA samples; +, PCR product from *Fusobacterium* DNA; -, template without DNA. (B) Melting curves of q-PCR amplicons obtained using *Fusobacterium*-specific primers in tumor samples. Blue lines, amplicons from tumor DNA samples; green line, amplicon from *Fusobacterium* DNA; red line, template without DNA; arrows, templates sequenced positively for *Fusobacterium*.

**Supplementary figure S3:** Effect of appendectomy on T-cell density after 3 DSS cycles (DSS-only protocol) and 1 week after surgery without DSS or AOM treatment (Surgery-only protocol).

Paraffin-embedded sections of the colon from each mouse after the DSS-only protocol in the appendectomy (n=15) and control (n=14 and not 15 due to a technical problem with one paraffin-embedded colon) groups were stained by immunohistochemistry (CD3 and CD8). (A) Representative CD3+ cell staining of a colon after appendectomy or sham laparotomy (control). (B) Quantification of CD3+ T-cell density in the lamina propria in the appendectomy and control groups after the DSS-only protocol. (C) Representative CD8+ cell staining of a colon after appendectomy or sham laparotomy (control). (D) Quantification of CD8+ T-cell density in the lamina propria in the appendectomy and control groups after the DSS-only protocol. Paraffin-embedded sections of colons from mice sacrificed one week after surgery (appendectomy, n=7 or sham laparotomy, n=7) without AOM or DSS treatment (Surgery-only protocol) were stained by immunohistochemistry (CD3 and CD8). (E) Representative CD3+ cell staining of a colon after appendectomy or sham laparotomy (control). (F) Quantification of CD3+ T-cell density in the lamina propria in the appendectomy and control group after the Surgery-only protocol. (G) Representative CD8+ cell staining of a colon after appendectomy or sham laparotomy (control). (H) Quantification of CD8+ T-cell density in the lamina propria in the appendectomy and control groups after the Surgery-only protocol. (I) Picture of an isolated lymphoid follicle in the colon. (J) Number of isolated lymphoid follicles in the colon in the appendectomy and control groups after the Surgery-only protocol. In all boxplots, the error bars represent the 25^th^, 50^th^ (median) and 75^th^ interquartile ranges. Comparisons of 2 groups were performed using the Mann-Whitney test with 2-tailed p-value. A p-value <0.05 was considered statistically significant.
