## Supplementary figures and images for "The appendix orchestrates T-cell mediated immunosurveillance in colitis-associated cancer"

### Supplementary figure S1

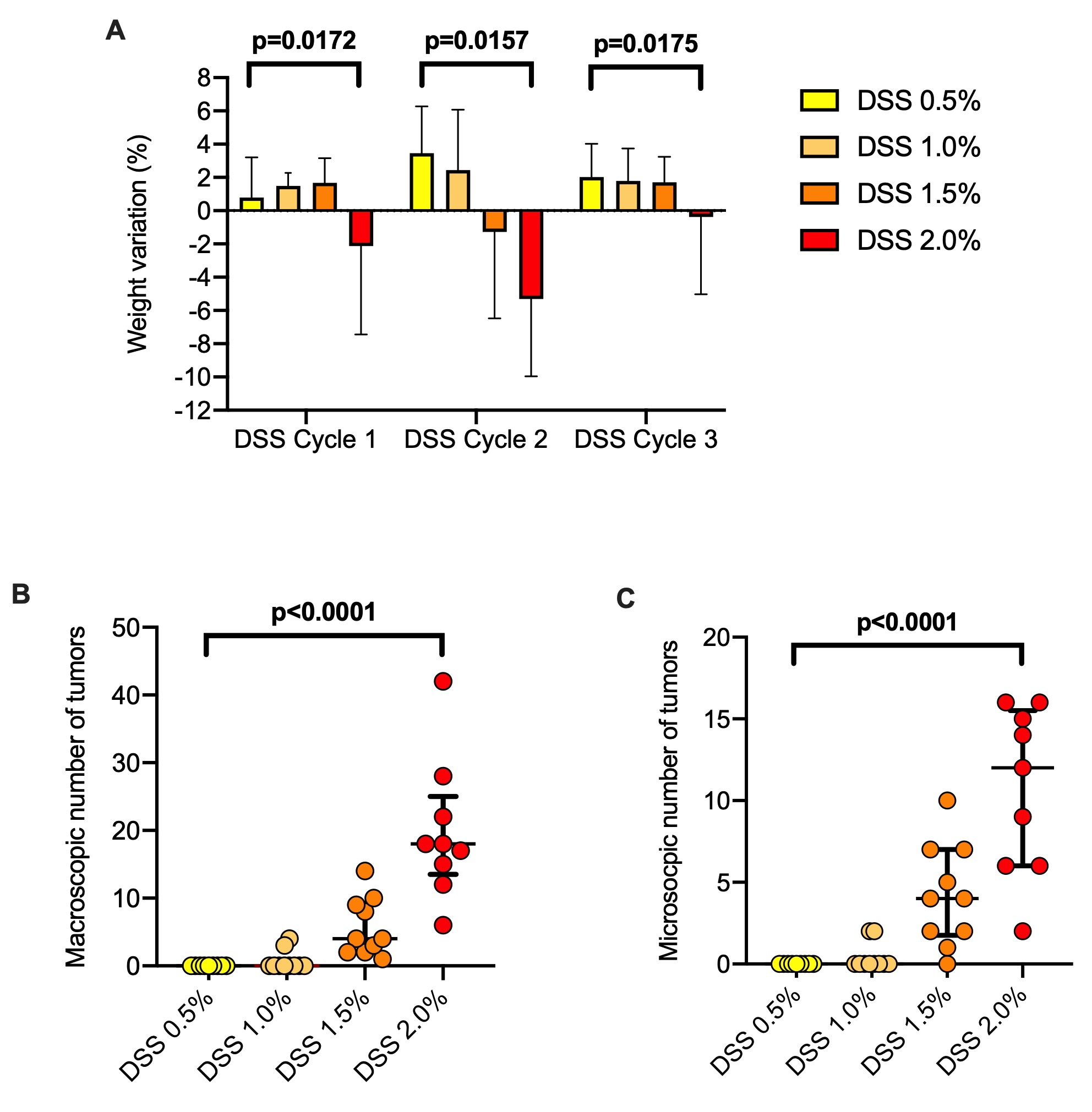

### Supplementary figure S2

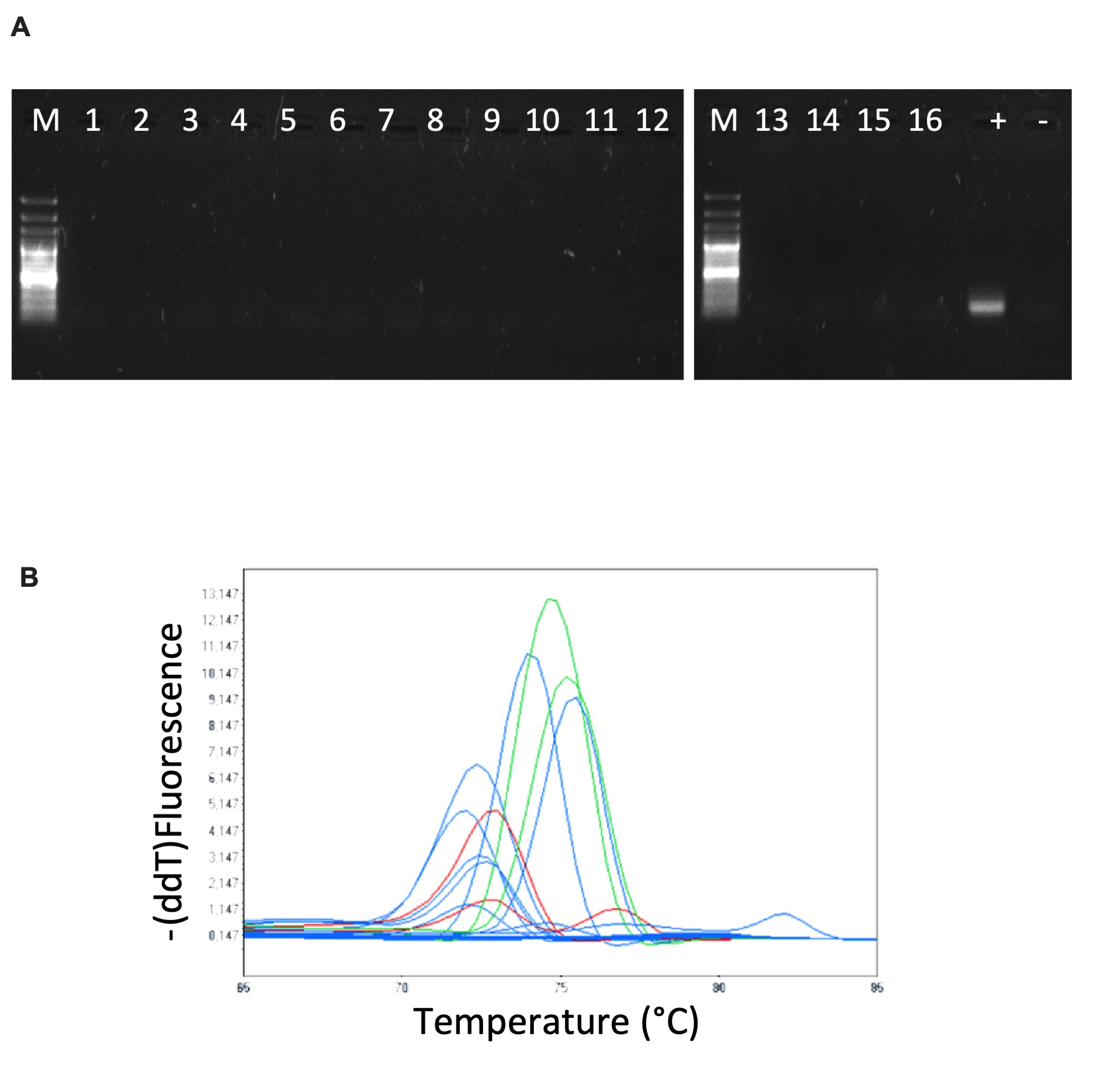

### Supplementary figure S3

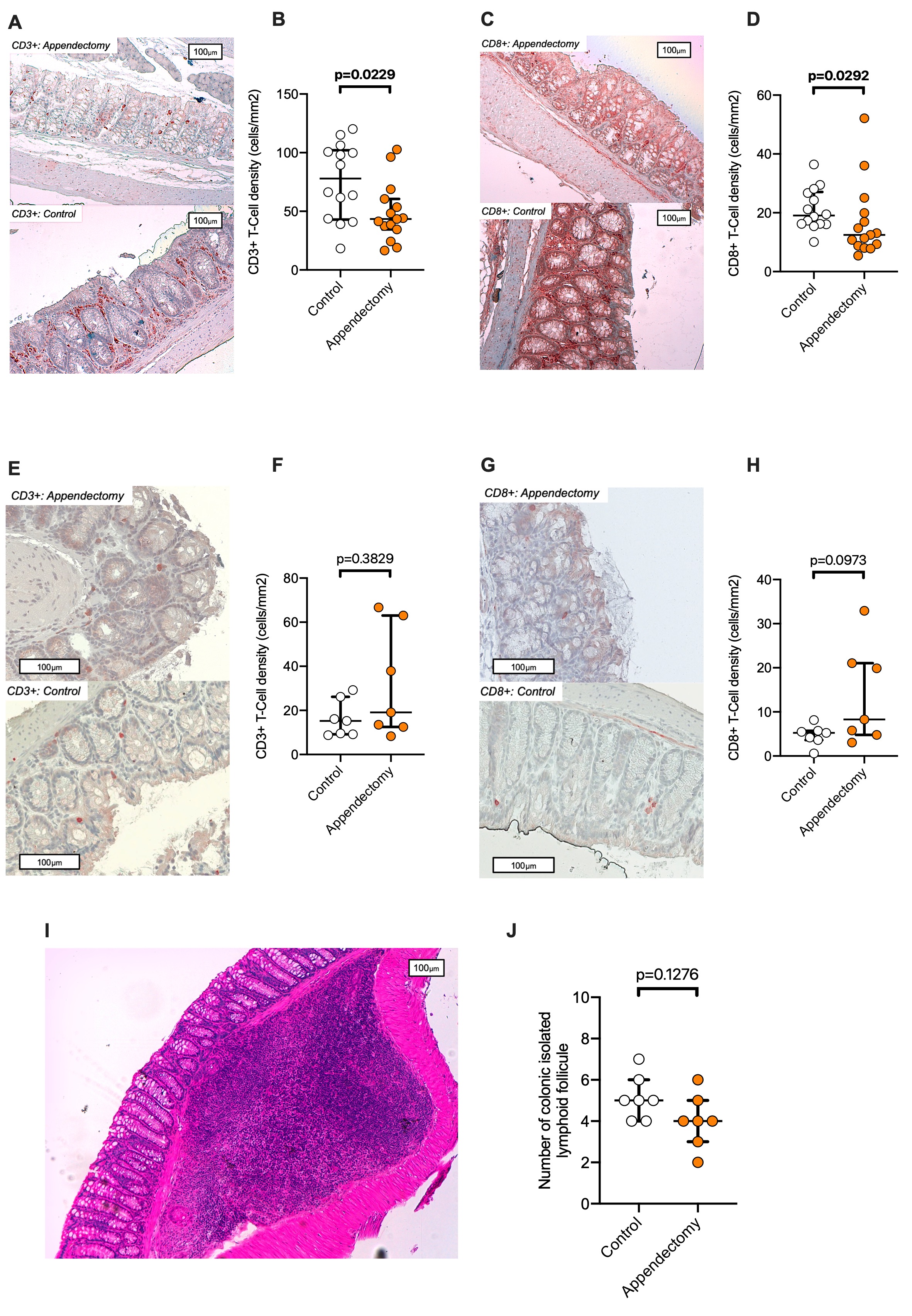
